# Genetic control of body weight by the human brain proteome

**DOI:** 10.1101/2022.02.11.22270813

**Authors:** Eloi Gagnon, Arnaud Girard, Émilie Gobeil, Jérôme Bourgault, Christian Couture, Patricia L. Mitchell, Claude Bouchard, Angelo Tremblay, Patrick Mathieu, Andréanne Michaud, Louis Pérusse, Benoit J. Arsenault

## Abstract

Genome-wide association studies (GWAS) have identified hundreds of genetic variants associated with body weight but the biological relevance of most remains unexplored. Given the critical role of the brain in multiple biological processes associated with body weight regulation, we set out to determine whether genetic variants linked with body mass index (BMI) could be mapped to brain proteins. Using genetic colocalization, we mapped 23 loci from the largest BMI GWAS (n=806,834) to brain proteins (obtained from a dataset of >7000 dorsolateral prefrontal cortex proteins measured by mass spectrometry in >400 individuals). We also performed a proteome-wide Mendelian randomization analysis followed by genetic colocalization, which allowed us to identify an additional 48 brain proteins linked with BMI. Multi-trait colocalization suggested that more than 75% of the protein quantitative trait loci (pQTL)-BMI associations are mediated via protein expression and not via RNA expression. Single-cell sequencing from the human brain cortex revealed that the genes expressing the proteins associated with BMI may be predominantly expressed in oligodendrocytes. In the Québec Family Study, a genetic risk score (GRS) including these brain pQTLs was associated with higher dietary carbohydrate intake and lower lipid intake whereas a GRS including the 67 variants most strongly associated with BMI was not associated with dietary intake. In conclusions, we identified 71 proteins expressed in the prefrontal cortex that may be critical regulators of body weight and possibly dietary intake in humans.

## INTRODUCTION

Identifying the genetic and biological determinants of body weight is critically important from an evolutionary and public health perspective. Over the past 40 years, changes in the food environment have undoubtedly contributed to the observed rapid increase in mean body mass index (BMI) in almost all countries. However, several lines of evidence suggest that genetic factors have shaped the individual response to the “obesogenic environment” as genetic susceptibility to an elevated body weight may be more penetrant in more modern environments ^1^. Twin and family-based studies revealed that genetic variations may account for 50–75% of BMI variance ^2,3^. Genome-wide association studies (GWAS) have identified hundreds of genetic loci influencing BMI ^4^. However, the biological function of the majority of these variants remains unexplored, highlighting the need for post-GWAS analyses. Using RNA-based technologies, many studies have suggested that most BMI-associated variants are located nearby genes enriched or exclusively expressed in the brain ^5,6^.

Studies using multi-omic technologies to facilitate the translation of genetic information into biological understanding of body weight regulation have been mostly limited to tissue RNA expression levels. Studying protein concentrations in addition to gene expression levels is critical, especially in light of recent studies describing a rather poor correlation between gene expression and protein levels, for instance in the brain ^7,8^. Since proteins are frequently the primary effector of biological function, their inclusion in post-GWAS analyses could improve our understanding of the neurobiological mechanisms influencing body weight regulation.

According to the prefrontal cortex model of obesity ^9^, genetic factors may influence food reward sensitivity and self-control, thereby influencing dietary self-regulatory abilities. The prefrontal cortex integrates appetite and satiety signals from the hypothalamus and the insula and could override these signals and suppressing the impulse to eat or planning and coordinating food acquisition. Neuroimaging studies have identified lower activation in response to a meal in a prefrontal cortex region, the left dorsolateral prefrontal cortex (DLPFC), in individuals with obesity compared to individuals without obesity ^10^. The DLPFC is a critical brain region involved in body weight regulation as it is associated with appetite control, food craving, food decisions, inhibition of eating and executive functioning (Gluck, Viswanath, and Stinson 2017). However, it is unknown if BMI genetic susceptibility loci influence brain protein concentrations, especially in the DLPFC or if proteins of this brain region are causally implicated in body weight control and/or dietary behaviour.

Here, we explore whether genetic variants associated with BMI could influence brain protein concentrations in a translational Mendelian randomization (MR) framework. We combine genome-wide data from 7,581 brain proteins measured in the DLPFC and BMI to map protein concentrations in the brain to known genetic variants associated with BMI. We also perform a brain proteome-wide MR (PWMR) study to identify brain proteins associated with body weight. We finally integrate the result of the mapping and PWMR approach to shed light on the biological mechanism of body weight control and dietary intake. The schematic overview of our analytical approach is presented in Fig. 1.

**Figure 1.**
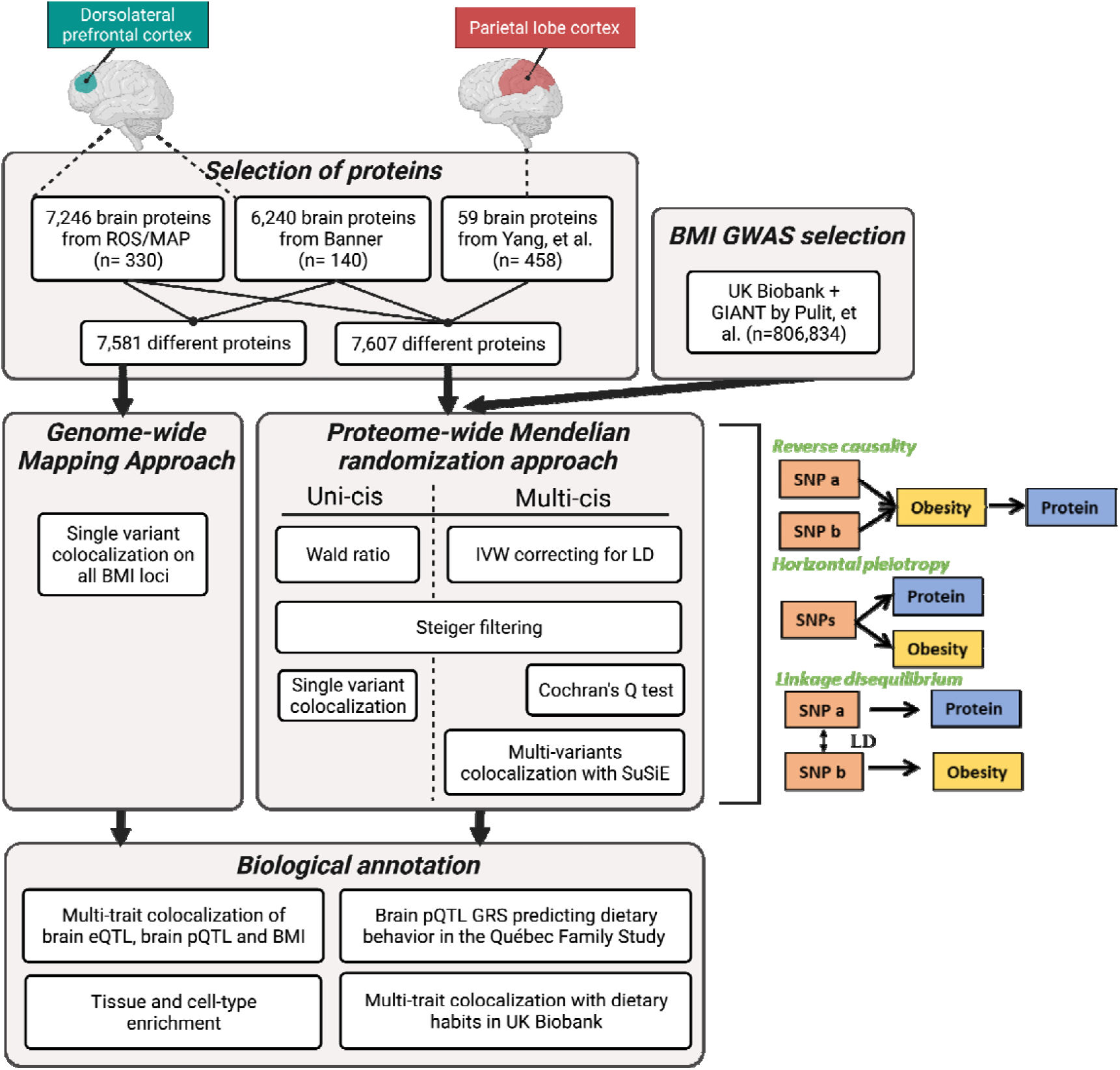
Schematic overview of the current study design. The study included a mapping approach, two types of proteome-wide Mendelian randomization approach, multi-trait colocalization analyses, a brain pQTL genetic risk score as well as tissue and cell-type enrichment analyses.

## RESULTS

### Genome-wide mapping of the body mass index susceptibility loci to the brain proteome

We extracted 543 genome-wide significant and independent (LD R2 < 0.001) BMI susceptibility loci from the largest BMI GWAS (806,834 participants of European ancestry) ^12^. We evaluated evidence of genetic colocalization between these variants and brain pQTLs from the Banner and ROS/MAP datasets ^7^. These two GWAS studies report on 7,581 brain protein levels measured by mass spectrometry from the DLPFC of 330 and 140 participants, respectively. Genetic colocalization is a Bayesian method evaluating the posterior probability of two traits sharing the same causal variant. Proteins with posterior probability of colocalization >0.80 were deemed to colocalize. Altogether, 23 BMI loci were mapped to brain proteins (Fig 2a and Supplementary Table 1).

**Figure 2.**
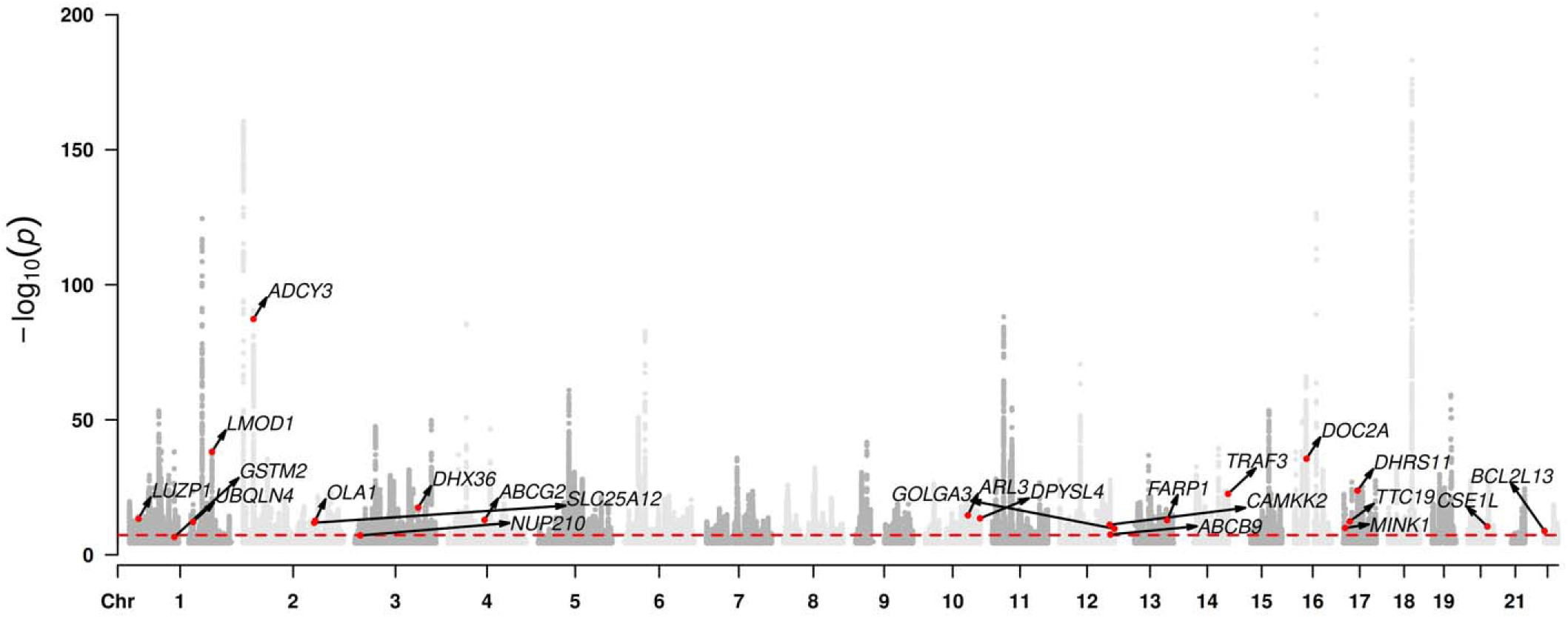
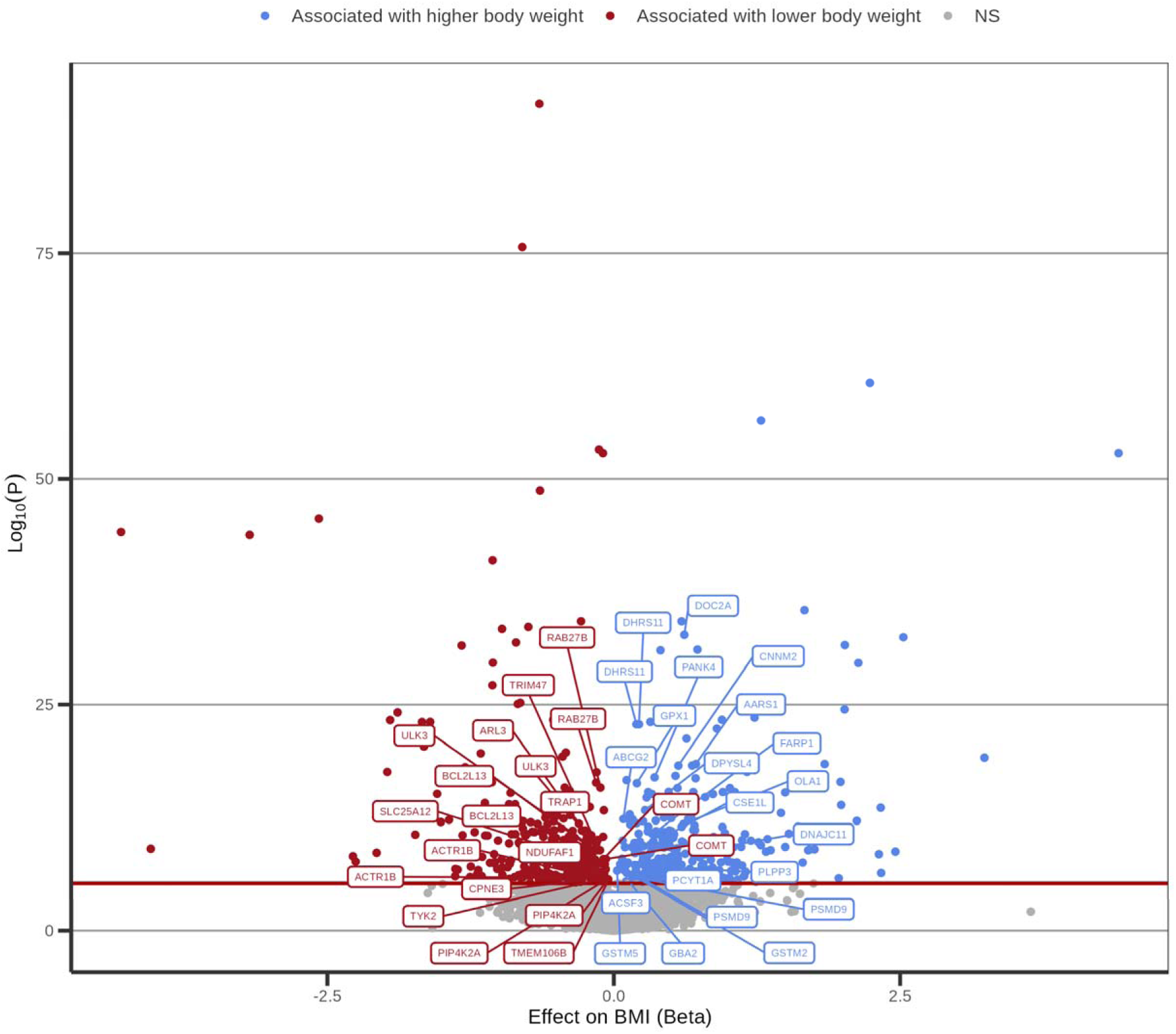
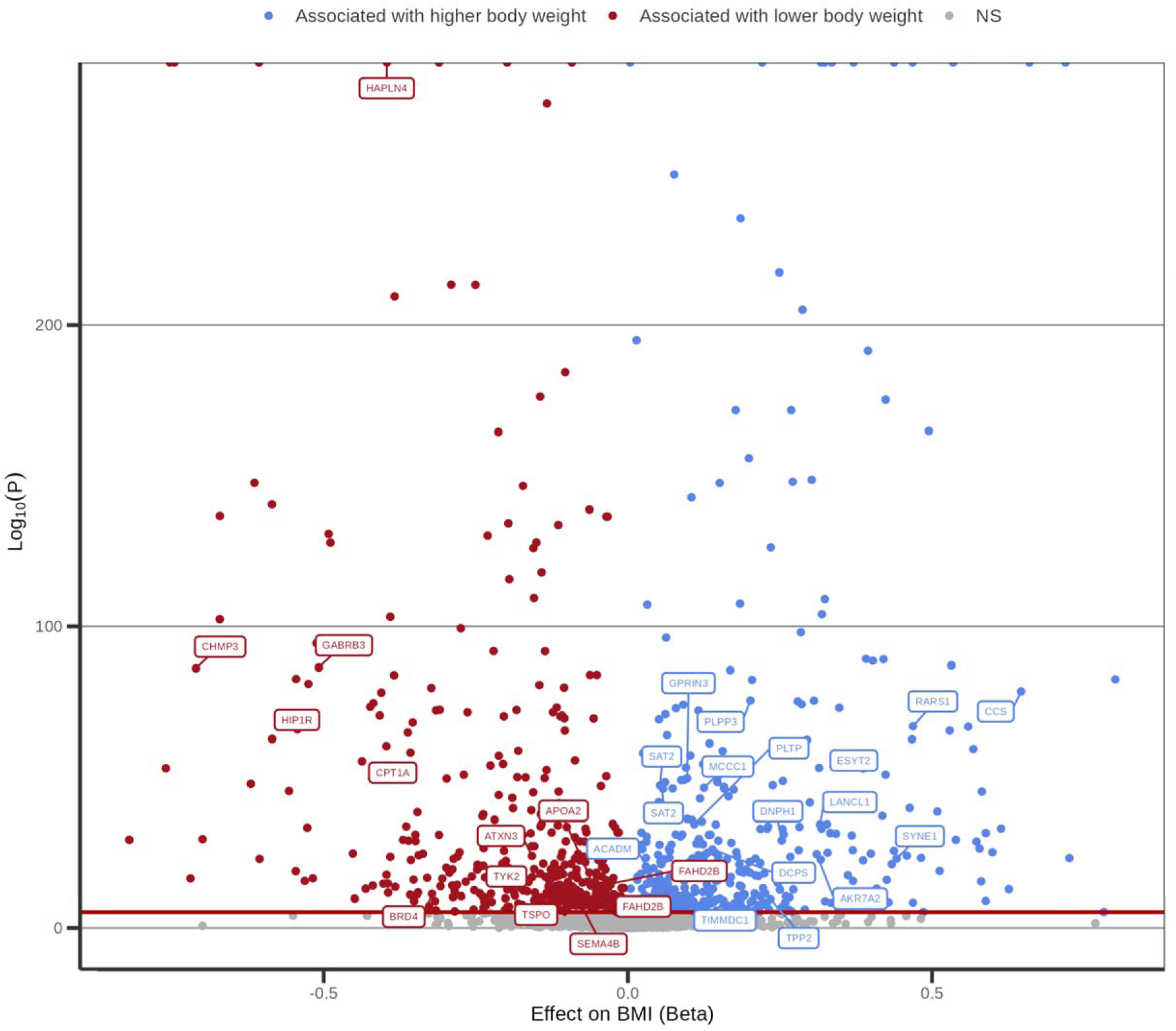
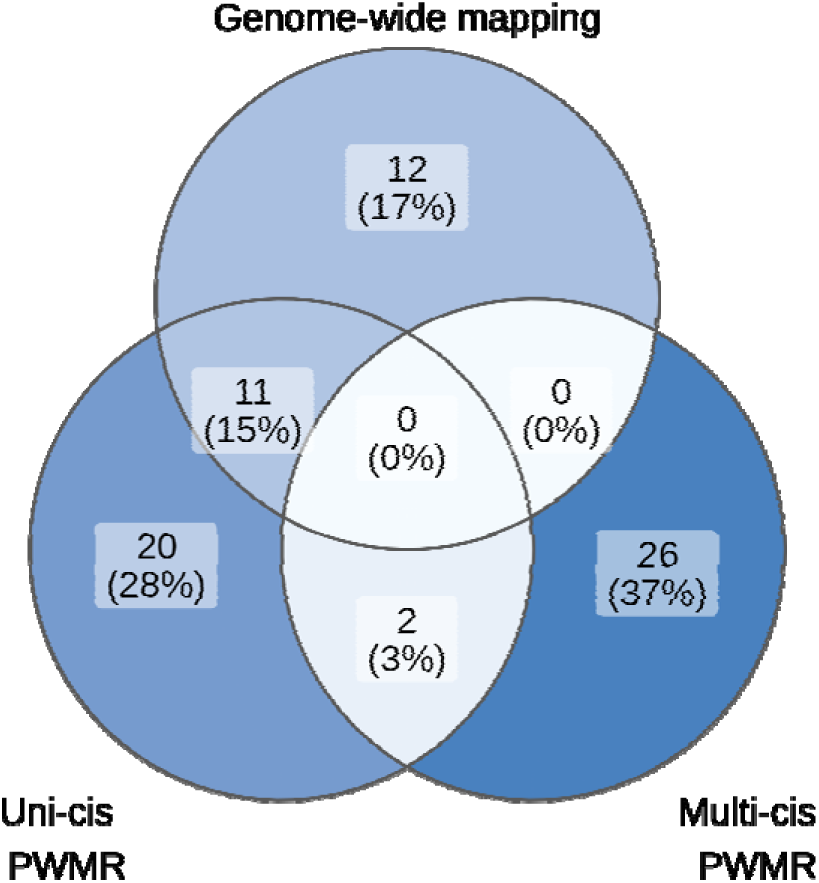
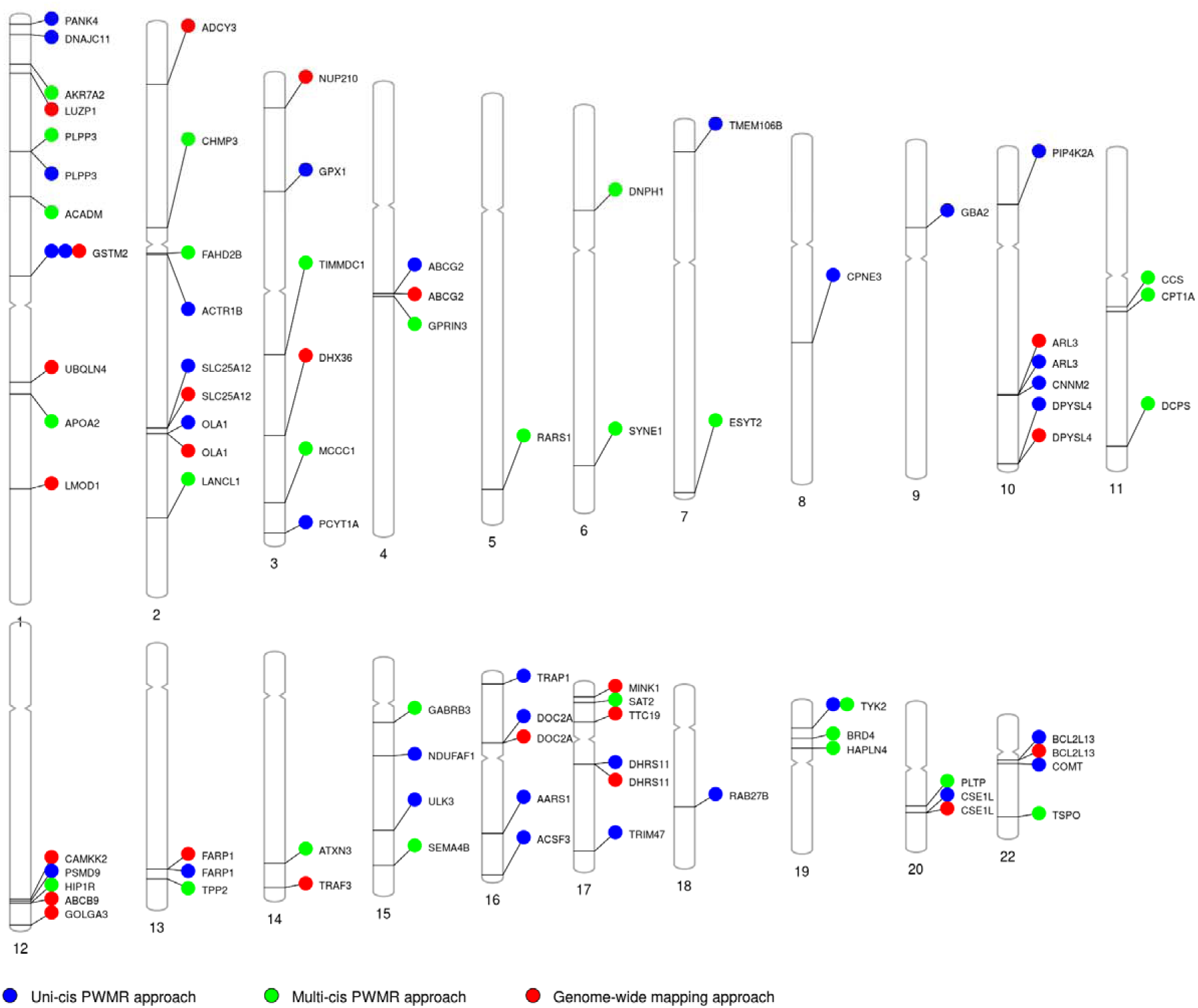
Brain proteins associated with the body mass index. A) Manhattan plot of body-mass index highlighting genetic regions with evidence of genetic colocalization with the human brain proteome. Genes highlighted on this plot include those with strong evidence of genetic colocalization (PPH4>0.80) B) Volcano plot representing the results of the brain proteome-wide Mendelian randomization study using the uni-cis approach. C) Volcano plot representing the results of the brain proteome-wide Mendelian randomization study using the multi-cis approach. D) Venn diagram reporting genetic regions with evidence of genetic colocalization with the humain brain proteome and the results of the brain proteome-wide Mendelian randomization study across the three methods. NS indicates not significant. E) PhenoGram representing the 71 identified proteins, their chromosomal location and the methods that identified them.

### Brain proteome-wide Mendelian randomization analysis of the body mass index

We used two strategies to determine whether brain proteins could be associated with BMI, an approach using one instrument only (uni-cis approach) and another one using more than one genetic instrument (multi-cis approach). In the uni-cis PWMR framework, we undertook two-sample MR and colocalization analysis to evaluate evidence for causal effects of 990 brain proteins on BMI (Fig. 2b). We evaluated proteins with at least one genome-wide significant (p<5e-8) cis-acting genetic instrument irrespective of their distance to BMI loci in the three brain pQTL datasets (Banner, ROS/MAP and Yang). We evaluated the effect of each brain protein on BMI using the Wald ratio method. We additionally performed colocalization analysis or, alternatively, linkage disequilibrium (LD) check (Methods) when colocalization was not feasible, to evaluate whether the brain proteins and BMI shared the same causal variant. A Bonferroni multiple testing correction was useed with the number of proteins (7607) as the number of tests. A total of 41 causal associations on 33 distinct proteins were identified (coloc PPH44 > 0.80 and p-value < 0.05/7607=6.6e-6) (Supplementary Table 2). No proteins in the Yang et al. ^8^ study passed the LD check. All associations exhibited evidence of protein to BMI directionality as tested with Steiger method. All associations had strong genetic instruments (F-statistic >10). All associations had genetic instruments located outside a 1 Mb window of the *MHC, APOE* and *ABO* gene regions, which are known to exhibit wide ranging pleiotropy. Results were homogenous when investigated separately in the UK Biobank and the GIANT BMI GWASes (Supplementary Table 3). Results also did not significantly differ when comparing Banner and ROS/MAP cohorts (Supplementary Table 4).

We next sought to identify additional brain proteins associated with BMI using a multi-cis MR analysis. This approach can yield more precise estimates (Gkatzionis et al., 2021). We fine mapped all genetic regions of brain proteins using the Sum of Single Effects (SuSiE) regression framework to prioritize multiple cis-acting causal variants ^14^. We used the selected variants as genetic instruments in an inverse variance-weighted MR analysis correcting for the LD matrix. We evaluated directionality with Steiger filtering and evaluated the heterogeneity of the estimates as a test for pleiotropy with Cochran’s Q test. Finally, we used colocalization analyses allowing for multiple causal variants to filter out linkage disequilibrium-contaminated associations using SuSiE ^15^. We defined evidence of genetic colocalization when all independent signals colocalized at PPH4 > 0.80. SuSiE finemapping revealed two likely causal variants for the Gamma-Aminobutyric Acid Type A Receptor Subunit Beta3 (GABRB3). GABRB3 is one the subunits of the receptor for gamma-aminobutyric acid, a major inhibitory neurotransmitter in humans. There was strong evidence of colocalization with BMI for both variants (0.99 and 0.99 respectively). The variants were in strong LD (R2=0.7) which can explain why colocalization with a single variant assumption failed. In total, 26 additional associations were identified (p-value < 0,05/7607; Cochran’s Q >0.05 (less heterogeneity than expected by chance); number of variants prioritized by SuSiE > 1; at least one instrument with p-value < 1e-3, all instruments Steiger filtering test p < 0.05 (no evidence for reverse causality), and all genetic instruments PPH4 > 0.8 (no evidence for bias by LD)) (Fig. 2c and Supplementary Table 5). Therefore, relaxing the single causal variant assumption allowed to find additional associations, where classic colocalization failed.

Fig 2d presents a Venn Diagram depicting the proteins identified by the genome-wide mapping, uni-cis and multi-cis approaches, respectively, and the overlap between these three methods that aimed at identifying brain proteins causally implicated in the regulation of body weight. Altogether, results of this analytical framework identified 71 brain proteins that may be causally implicated in the regulation of body weight in humans. Fig 2e presents the chromosomal location of these proteins.

### Brain expression levels of genes encoding body weight-regulating brain proteins

Many studies pinpointing the brain’s role in the genetic regulation of body weight have relied on RNA-based methods ^5,6^. However, Yang et al. ^8^ revealed that only 20% of genetic variants affecting protein levels affect also brain RNA levels in the brain. Therefore, we performed multi-trait colocalization analysis to evaluate whether gene expression lays in the causal pathway linking brain pQTLs to BMI. As multi-trait colocalization does not allow for multiple causal variants, we included proteins identified in the mapping and uni-cis PWMR approach but not those identified from the multi-cis PWMR approach. We performed multi-trait genetic colocalization using the gene expression levels in the Broadman area 9 from GTEx, the pQTL from the DLPFC and BMI. The Broadman area 9 was chosen because it contributes to the region of the brain where protein expression was quantified (the DLPFC). This analysis revealed that a minority (11/45) of proteins had evidence of multi-trait genetic colocalization with BMI and gene expression levels (Supplementary Table 6). Fig 3 illustrates this concept by depicting multi-trait genetic colocalization at two genetic loci, *ADCY3* (adenylate cyclase type 3) and *DOC2A* (double C2-like domain-containing protein alpha). These genes were selected to highlight a difference in multi-trait genetic colocalization with gene expression levels. *ADCY3* had a posterior probability of multi-trait colocalization of 0.94 (Fig. 3a), meaning that the effect of the ADCY3 protein levels on BMI is consistent with higher gene expression levels. In contrast, no evidence of genetic colocalization with brain protein and gene expression levels were found in the case of DOC2A. This absence of genetic colocalization between brain protein and gene expression levels was found for the majority of body weight-regulating brain proteins. These results therefore suggest that relying solely on gene expression levels to identify novel brain factors associated with body weight regulation might be insufficient.

**Figure 3.**
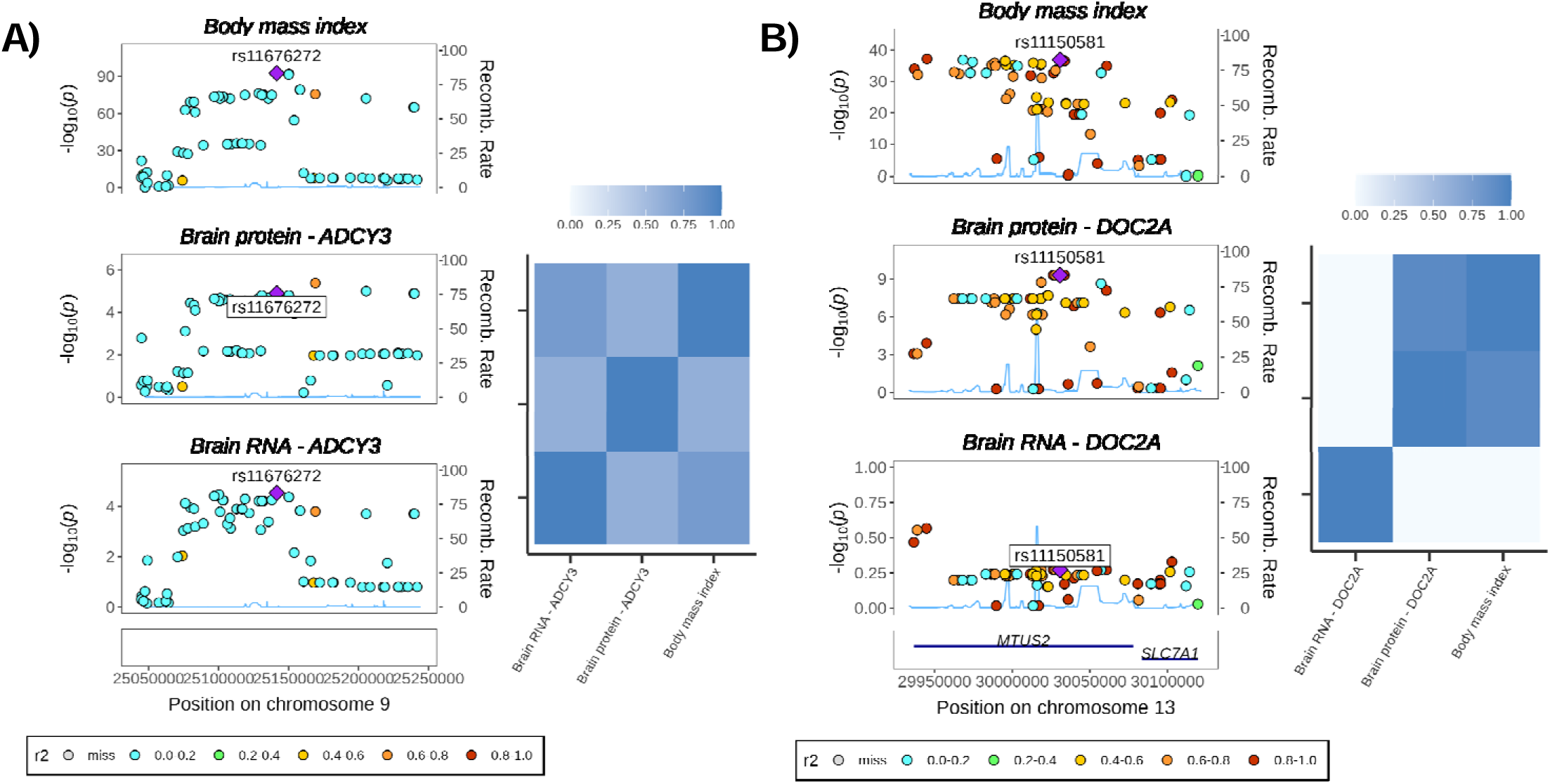
Mutli-trait colocalization for RNA levels, protein levels and BMI. A) ADCY3 as an example of multi-trait colocalization between brain RNA, brain protein and BMI. B) DOC2A as an example of multi-trait colocalization between protein levels and BMI, but not brain RNA levels.

### Tissue and cell specificity of the body weight-regulating brain protein

Having identified 71 candidate brain proteins linked with BMI, we next sought to identify the tissue and cell-type where these genes are preferentially expressed. We first evaluated brain tissue specificity for the 71 causal proteins using RNA expression from the GTEx-V7 database. We obtained the tissue-specific gene expression metric (tau). Genes with evidence of tissue-specific expression have a tau value closer to 1 while ubiquitous genes have a tau value closer to 0. The majority of genes were expressed ubiquitously across tissue (65/71 tau < 0.7) (Supplementary Table 7). Among genes with tissue specific expression (tau > 0.7) 4/6 had evidence of higher expression in the brain (*HAPLN4, GPRIN3, GABRB3* and *DOC2A*). We then performed tissue and cell-type enrichment analyses to test the hypothesis that the 71 causal proteins were enriched in specific tissue and specific cell-types. Tissue expression enrichment using MAGMA software ^16^ revealed that the genes identified are preferentially expressed in key brain regions involved in the control of appetite (hypothalamus), reward (amygdala and hippocampus) and inhibition (brain frontal cortex and anterior cingulate cortex) (Fig.4-A). The genes are enriched exclusively in brain tissues. Cell-type enrichment analysis using FUMA ^17^ and single-cell RNA sequencing of 333 cells from the human cortex revealed that the 71 candidate causal genes are preferentially expressed in oligodendrocytes (Fig. 4-B).

**Figure 4.**
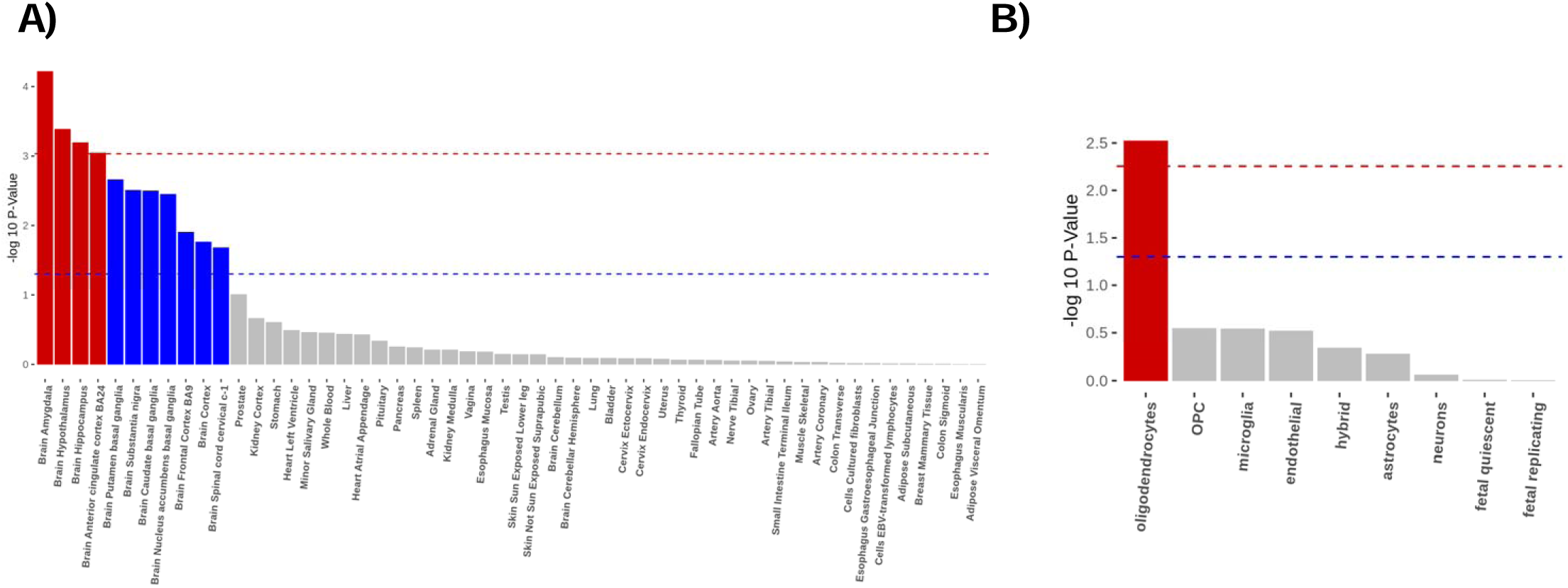
Tissue and cell-type enrichment of the 71 causal genes. A) MAGMA tissue enrichment analysis using GTEx RNA-seq data on 53 specific tissue types. B) FUMA’s cell-type enrichment analysis in the cerebral cortex. The red dashed line is the significance threshold with multiple testing correction and red bars pass this threshold. The blue dashed line is the significance threshold at 0.05 and blue bars pass this threshold. Grey bars are tissues that display no enrichment.

### Eating and dietary traits linked with brain protein quantitative trait loci

In order to determine how brain pQTLs may influence body weight, we investigated the associations between a brain pQTL genetic risk score (GRS) and eating and dietary traits in 750 participants of the Québec Family Study (QFS). In this study, eating behavioral traits were assessed using the three-factor eating questionnaire and dietary intakes were assessed using 3-day dietary records. The brain pQTL GRS was built using all independent lead SNPs of the 71 candidate causal genes and its predictive value was evaluated with linear regressions correcting for age, sex, and under and over-reporter status of total energy intake. Each SD increase in brain pQTL GRS was associated with an increase in the percentage of carbohydrate intake (1.04% 95% CI=0.54-1.55, p=5.0e-05) and a decrease in the percentage of lipid intake (−0.74% 95% CI=-1.19-0.29, p=1.3e-03) (Fig. 5-left panel). By comparison, a GRS calculated with 67 BMI lead SNPs had no effect on the macronutrient composition of the diet (Fig. 5-right panel), but was associated with eating behaviors such as susceptibility to hunger and disinhibition (Supplementary Table 8b) as previously described ^18^. This providing support for the specificity of the brain pQTL GRS as a determinant of dietary intake and especially with high carbohydrate intake (Supplementary Table 8a).

**Figure 5.**
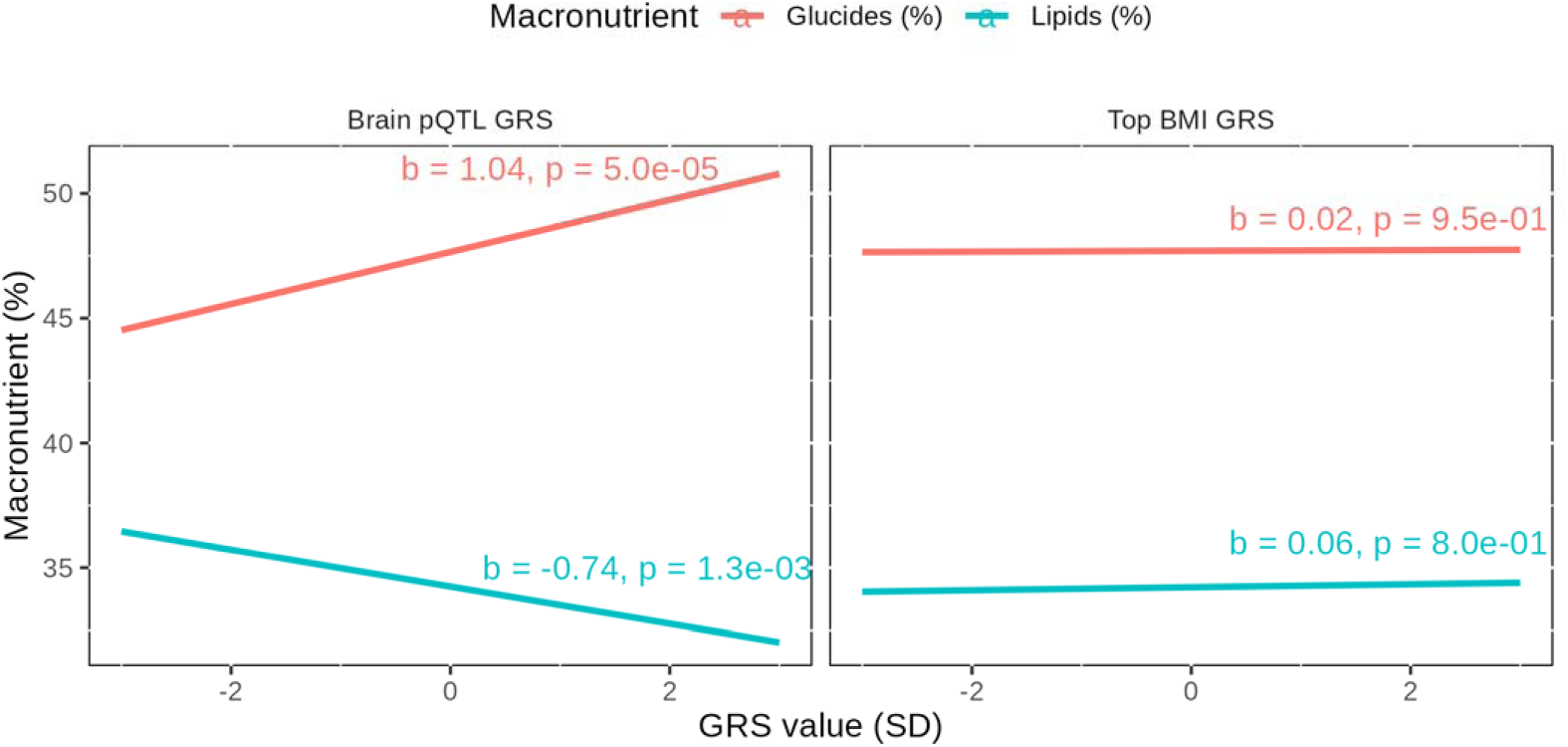
Association between a genetic risk score (GRS) based on brain protein quantitative trait loci (pQTL) linked with the body mass index (BMI) and a GRS based on the most significant BMI susceptibility loci on macronutrient intake in the Québec Family Study. Left panel) Impact of the brain pQTL GRS on carbohydrate and lipids intake. Right panel) Impact of the top BMI GRS effect on carbohydrate and lipids intake.

### Shared genetic etiology between brain protein quantitative trait loci and dietary habits in the UK Biobank

Having established the association between brain protein expression with higher carbohydrate intake, we next sought to establish which specific proteins could be linked with dietary habits. Colocalization analysis between all causal proteins and 63 dietary habits derived in participants of the UK Biobank ^19^ revealed dietary habits as potential mediator between certain brain protein and BMI (Fig. 6). This analysis identified 11 proteins sharing genetic etiology with at least one dietary habits (Supplementary Table 9). When proteins colocalized with more than one dietary habit, multi-trait colocalization identified 4 proteins affecting clusters of dietary habits. Notably, DOC2A and ADCY3 had a multi-trait colocalization with a cluster of five and ten dietary habits respectively. For six proteins, ACTR1B, ADCY3, BCL2L13, GBA2, RAB27B and ULK3, >60% of the posterior probability of colocalization of the primary genetic association with BMI, dietary habits and the respective protein levels were explained by a single variant (rs11692435, rs1018218, rs11538, rs3750434, rs8085272 and rs936227 respectively).

**Figure 6.**
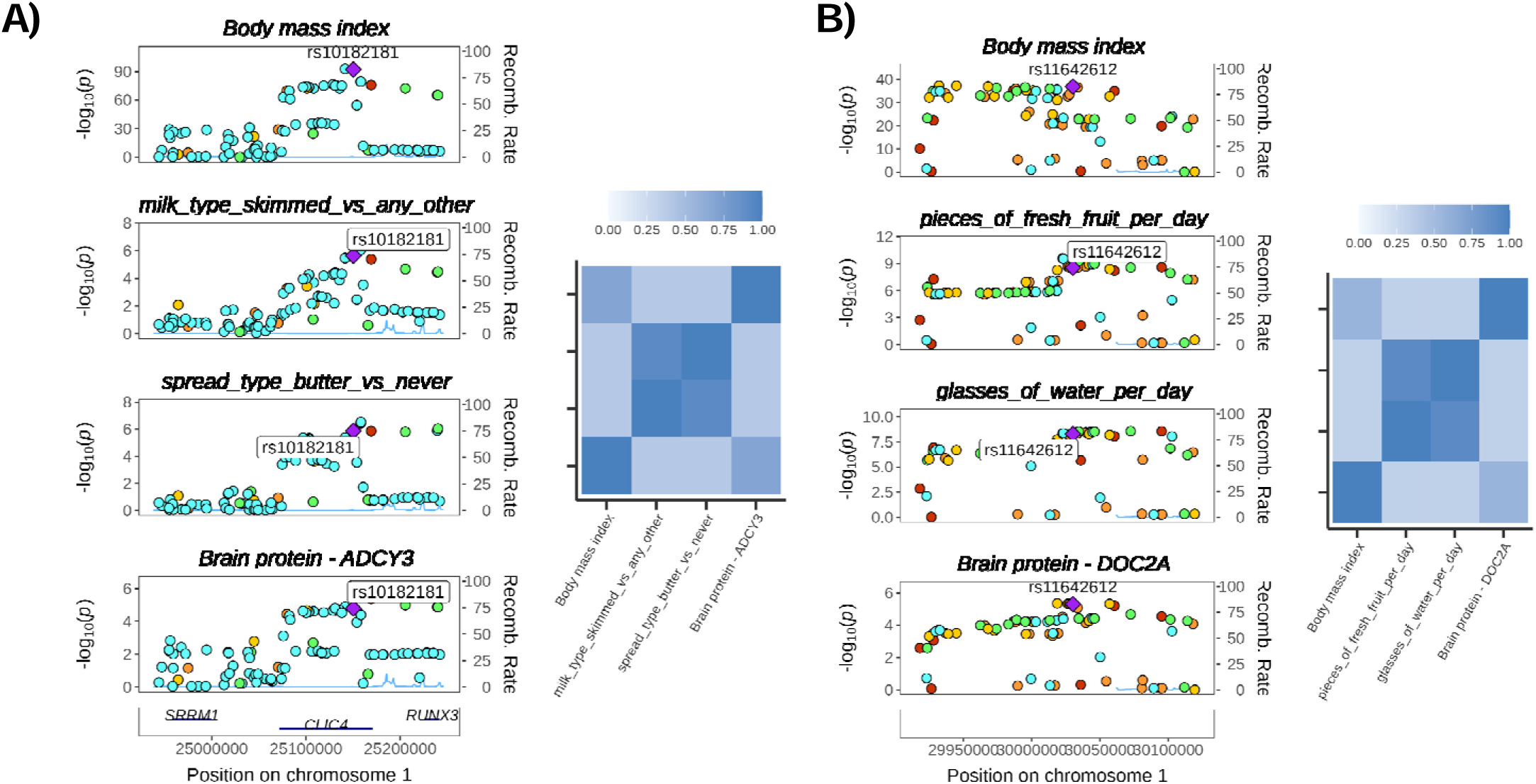
Multi-trait colocalization for brain protein levels, dietary habits, and body mass index in the UK Biobank. Stack association plot depicting the association of the 100Kb upstream and downstream at the A) *ADCY3* and B) *DOC2A* loci. The annotated genetic variants are the prioritized causal variants by HyPrColoc. Prior sensitivity heatmaps are also presented. We iteratively varied choice of priors to verify the stability of the cluster as a sensitivity analysis. Values closer to one are less sensitive to variation of priors.

## DISCUSSION

Given the critical role of the brain in body weight regulation, we set out to explore whether genetic variants associated with BMI could be linked to brain protein concentrations. Using a combination of genetic colocalization and Mendelian randomization analyses, we identified 71 brain proteins that may be involved in the control of body weight. About three quarter of the pQTLs affected BMI via protein levels and not via RNA levels. Although the majority of the genes encoding the 71 brain proteins are ubiquitously expressed, the expression of these genes appears to be enriched in brain regions involved in the regulation of body weight and further enriched in oligodendrocytes within the cortex. Presumably, part of the effect of these proteins on body weight is mediated by dietary habits as shown by multi-trait colocalization with dietary intake in the UK Biobank and the Québec Family Study. Altogether, these results support that brain proteins may play a central role in regulating body weight and identify plausible biological and behavioral mechanisms implicating the human brain proteome and dietary habits in the regulation of body weight.

While most of the proteins that we identified are potentially new candidate genes regulating body weight, some of them had already been identified. First, *ADCY3* was identified concurrently by three different studies as a susceptibility locus for obesity ^20–22^. Second, DOC2A is a brain specific protein involved in Ca(2+)-dependent neurotransmitter release located in the 16p11.2 gene region. The number of genomic copies in this region is inversely correlated with body mass index (BMI) and a deletion of this locus has been associated with a highly penetrant form of obesity ^23^. Third, the catechol-O-methyltransferase (*COMT)* enzyme breaks down dopamine terminating its function in the prefrontal cortex. Individuals with low COMT activity are more likely to report binge eating behaviors ^24^ and to have higher BMI ^25^. Finally, CaMKK2 expression is positively associated with BMI and food intake in rodents. Acute pharmacologic inhibition of CaMKK2 in wild-type mice, but not CaMKK2 null mice, inhibits appetite and promotes weight loss ^26^. The biological significance of the identified proteins and the mechanisms by which they contribute to the regulation of body weight require further investigation.

In this regard, follow up analyses performed on the 71 proteins identified allowed us to confirm plausible biological mechanisms of body weight regulation. Using multi-trait colocalization analyses, we showed that a large proportion of brain pQTL-BMI associations are not mediated by differences in gene expression levels. This finding is in line with recent evidence showing that a large proportion of pQTL are not eQTL in the brain ^7,8^. We showed that the 71 candidate genes are enriched in brain regions involved in appetite control such as the amygdala, hypothalamus, hippocampus, and anterior cingulate cortex. Similarly, RNA-based studies have shown that genes located nearby BMI loci are primarily expressed in the brain ^6^. Using single-cell sequencing in the brain cortex, we provide evidence that these 71 genes are enriched in oligodendrocytes. The main function of this cell type is to support axon myelination in the central nervous system. Although the role of oligodendrocytes in the regulation of body weight has yet to be studied in depth, a recent cross-sectional study reported differential patterns of myelination in individuals with versus without obesity ^27^. A novel finding is the potential role for some of the identified proteins in influencing food choices specifically high carbohydrate intake. How proteins expressed in the human prefrontal cortex specifically drive different food choices will need to be further explored in functional and interventional studies.

Our results extend those recently reported from a similar PWMR study focused on neurological disorders. Wingo et al. recently performed a proteome-wide association study on depression and used BMI as negative control ^28^. Their PWMR analysis revealed 217 candidate proteins linked with BMI. However, genetic colocalization analyses were not performed so the authors could not rule out confounding by LD. Confounding by LD occurs when the top associated *cis*-pQTL is in LD with two distinct causal variants, one affecting protein expression and the other affecting trait variation. Recent evidence have suggested that even when a protein-disease association has strong evidence from MR (p<5e-6), up to 38% of the associations do not have evidence from colocalization ^29^. This highlights the need for colocalization analyses in preventing confounding by LD for PWMR analyses. Using an analytical framework that combines genome-wide mapping of known BMI loci, uni- and multi-cis PWMR and genetic colocalization analyses, we replicated 30 of Wingo et al. hits and further identify 41 new hits. Only a minority (49/217) of the proteins identified by Wingo et al., colocalized with BMI. The reported brain pQTLs associated with BMI in our study are therefore less prone to confounding by LD and our list of proteins is more conservative and more likely to be causally linked with BMI.

Our approach has limitations that we believe may have led to an underestimation of the impact of the brain proteome on body weight control. First, although 7606 proteins were measured, the small sample size of the brain pQTL datasets led to only 703 proteins with a genome-wide significant pQTL available for analysis. Second, analysis was limited to the DLPFC and parietal lobe cortex as protein expression GWAS of other relevant brain regions, such as the hypothalamus or the insula, have yet to be performed. Third, due to the small sample size of the GWAS on proteins, only common variants (MAF > 0.05) were included, although it is known that BMI is also influenced by less frequent and rare variants ^30^.

The heritability of BMI has been estimated to reach 50-75% and several hundred genetic loci have been suggested to contribute to the differences in body weight across individuals. This study identifies the human brain proteome as a potentially important contributor to the heritability of BMI and lend support to a potential interaction between the human brain proteome and the evolving food environment that may influence food preferences such as sugar intake. Taken together, the results of this study suggest that inter-individual differences in the human brain proteome might explain to a certain extent why food preferences and food choices vary considerably from one individual to another and why the genetic susceptibility to an elevated body weight may be more phenotypically expressed in the modern food environment.

## METHODS

### Study populations

#### Main study exposures

We performed colocalization analyses and two-sample Mendelian randomization investigations using GWAS summary-level data. There was no sample overlap between data sets. All participants were of European ancestry. A total of three brain pQTL datasets were used 1) **ROS/MAP** dataset: We extracted brain pQTL from a GWAS on protein concentrations in the dorsolateral prefrontal cortex of 330 older adults (age median 89 range [71–106.5]) ^7^. Participants were drawn from the Religious Orders Study (ROS) and Rush Memory and Aging Project (MAP) cohorts. The ROS and MAP Project are ongoing longitudinal clinical-pathologic cohort studies ^31^. Both studies recruited participants without known dementia. Proteomic profiling was performed using liquid chromatography coupled to mass spectrometry and isobaric tandem mass tag peptide labelling. Genotyping was performed by whole genome sequencing or genome-wide genotyping by either Illumina OmniQuad Express or Affymetrix GeneChip 6.0 platforms. Reads were aligned to the GRCh37 human reference panel. Brain pQTL were derived using linear regression adjusted for age at death, sex, batch, post-mortem interval, dementia status at death and first ten principal components; 2) **Banner dataset**. We extracted brain pQTL from a GWAS on protein concentrations in the dorsolateral prefrontal cortex of 140 older adults (median age 86 range [66–103]) ^7^. Participants were drawn from the Banner Sun Health Brain and Body Donation Program (Banner). Banner has banked more than 1600 brains from cognitively normal volunteers in Phoenix, Arizona. Proteomic profiling was performed with a similar approach as described for ROS/MAP. Genotyping was performed with Affymetrix Precision Medicine Array. Reads were aligned to the GRCh37 human reference panel. Brain pQTL were derived using linear regression adjusted for age at death, sex, batch, dementia status at death, post-mortem interval and first ten principal components. 3) **Yang dataset**. We extracted brain pQTL from a GWAS on protein concentrations in the parietal lobe cortex of 458 participants of European ancestry from the Washington University ^8^. Among the 458 participants, there were 345 with Alzheimer’s disease, 12 cognitively normal controls and 102 with unknown or other statuses (e.g., dementia or other neurological diseases). The age was normally distributed with a mean of 83.3 years and standard deviation of 10 years (57% women). In this study, there was an extremely high correlation (Pearson’s r >0.98), between the pQTL of Alzheimer cases and healthy cases indicating that the association of the genetic variants with protein levels does not depend on disease status. Proteomic profiling was performed with a multiplexed, aptamer-based approach to measure the relative concentrations of proteins. After QC, 1079 proteins and 380 samples were kept. Samples were genotyped on multiple genotyping platforms from Illumina. Reads were aligned to hg19/GRCh37. Genome-wide summary statistics were not available in this dataset. Only genome wide significant pQTLs (p<5e-8) were available when these analyses were conducted.

#### Main study outcome

We used GWAS summary statistics from a meta-analysis of the Genetic Investigation of Anthropometric Traits (GIANT) consortium and the UK Biobank totalling 806,834 participants of European ancestry ^12^. Measures of BMI were self-reported, measured in a laboratory or measured in a healthcare setting. Measures were corrected for age, age squared, sex, principal components and study site. The resulting residuals were transformed to approximate normality with SD of 1 using inverse normal scores.

### Statistical Analyses

#### Brain protein quantitative trait loci mapping of body mass index

We performed genome-wide mapping of BMI susceptibility loci to the brain proteins of the Banner and ROS/MAP datasets. We included genes that were at +/- 100Kb from one of 543 independent (LD R2 = 0.001) and genome wide significant top hits. We performed genetic colocalization analysis with a single causal variant assumption using the *coloc* R package with default priors. Proteins with posterior probability of colocalization over 0.80 were deemed as colocalized signals, meaning that the protein and phenotypes likely share the same causal variant.

#### Brain proteome-wide Mendelian randomization analysis of the body mass index

We performed single SNP MR using the genome-wide significant cis-acting pQTL with the lowest p-value. MR estimate on each pQTL and BMI association were obtained with the Wald ratio. The Wald ratio is calculated by dividing the SNP-outcome effect by the SNP-exposure effect. We also used a multi-cis MR approach where we selected multiple independent cis-acting variants as genetic instruments. Multi-cis MR analysis, that is, using multiple genetic variants from a single locus even if the variants are correlated can result in more precise estimates (Gkatzionis et al., 2021). For this analysis, we included as genetic instruments all cis-acting SNP prioritized by fine mapping with SuSiE. We used a generalized inverse-variance weighted model taking into account the LD correlation matrix between the multiple cis-acting instrument ^32^. We used the *MendelianRandomization* R package to calculate the MR estimates ^33^ and obtained the correlation matrix from the 1000 Genomes European ancestry reference samples. To assess instrument strength, we used the F-statistic ^34^, and to quantify the variance explained, we used the R2 value ^35^. We corrected for multiple testing with a Bonferroni procedure using the number of different proteins as the number of tests.

#### Sensitivity analyses of the proteome-wide Mendelian randomization studies

We performed a range of sensitivity analyses on the associations that survived the multiple testing correction procedure to remove spurious associations. Spurious association can occur because of reverse causality, horizontal pleiotropy, or linkage disequilibrium. We assessed bias by reverse causality with Steiger filtering, horizontal pleiotropy with Cochran’s Q tests and linkage disequilibrium with colocalization analyses. First, to distinguish causality from reverse causality, we used Steiger filtering ^36^. The Steiger filtering test tags variants when their association with the outcome is stronger than with the exposure ^37^. The test was implemented in the *TwoSampleMR* R package. Genetic instruments with nominally significant test (p <0.05) were kept as it indicated evidence for an exposure to outcome direction of association. Second, to evaluate if the associations were due to horizontal pleiotropy, we performed Cochran’s Q test. Excluding horizontal pleiotropy is challenging, but the use of multiple instruments can indirectly address the issue. Classically employed robust MR methods (MR Egger regression, Weighted Median, etc.) require at least three independent genetic instruments. Due to the low number of independent pQTLs, performing these sensitivity analyses was not possible. For multi-SNP MR analyses, we performed instead Cochran’s Q test to evaluate the heterogeneity of the estimates ^38^. Cochran’s Q evaluates the null hypothesis that all genetic variants estimate the same causal parameter. When the null was rejected at p-value <0.05, the association was categorized as pleiotropic. Third, to evaluate bias due to linkage disequilibrium, we performed a series of genetic colocalization analyses. We evaluated the posterior probability that both the protein and the BMI shared a single variant using a Bayesian model implemented in *coloc* R package ^39^. We used the default priors for the analysis. We used a posterior probability H4 > 0.80 as a threshold to suggest that the two associations shared the same causal variant within the pQTL cis region. We also performed colocalization relaxing the single causal assumption to evaluate if associations showed evidence of colocalization at multiple loci. Classical colocalization makes the assumption that only one causal variant is shared between traits. However, multiple causal variants located in close proximity can influence protein expression. SuSiE regression framework is an approach that allows the evaluation of multiple causal variants simultaneously. We evaluated all SNPs in a 200 kb window of the gene and interpreted posterior probability H4 > 0.80 as evidence for colocalization. The Yang dataset reported only pQTLs with a p-value <5e-8, therefore not allowing us to perform colocalization analyses due to insufficient genetic coverage. For pQTL analyses in the Yang study, we conducted a LD check instead ^29^. We estimated if the genetic instrument proxying protein levels were in LD with the 30 more robust BMI SNPs in a 1Mb window of the genetic instrument. When any of the 30 BMI genetic variants were in strong LD (R2> 0.8) with the sentinel variant of the Yang study, we considered the two studies to share the same causal variant.

#### Multi-trait colocalization with gene expression levels

We investigated shared genetic etiology across brain eQTL, brain pQTL and BMI using multi-trait colocalization analyses with the HyPrColoc R package ^40^. GWAS summary statistics of brain RNA expression was derived from the Broadman area 9, which was available for 156 participants from the Genotype-Tissue Expression (GTEx-V8) database. HyPrColoc extends the coloc methodology by estimating posterior probability of more than two traits sharing the same causal variants. If the traits do not share a causal SNP, a branch-and-bound selection algorithm is employed to discover subsets of traits that colocalize. We used the default uniform prior for the analysis. Prior.1 (the prior probability of a SNP being associated with one trait) was set to 1e-4 and prior.c (the probability of a SNP being associated with an additional trait given that the SNP is associated with at least one other trait) was set to 0.02. Colocalization analyses are sensitive to the choice of prior. We therefore iteratively varied the choices of prior to assess the stability of clusters as a sensitivity analysis. The colocalization events were visualized with regional stack lot created with the *gassocplot* package.

#### Tissue specificity, tissue enrichment and cell-type enrichment

We evaluated tissue specificity, tissue enrichment and cell-type enrichment for the causal proteins. First, we evaluated tissue specificity of the genes expressing these proteins by calculating transcripts per million (tpm) from all available tissues in GTEx-V7 except sex-specific tissue and tissue from the urinary system. All tissues with genes tpm <1 were categorized as not expressing the gene. We then calculated tau as a measure of tissue specificity (Yanai et al., 2005). The values of tau ranges from 0 to 1, where zero indicates a ubiquitous gene expression and one indicates specific gene expression. It has been shown that tau is the best choice for calculating tissue specificity ^42^. Second, we performed tissue enrichment analysis using MAGMA ^16^. Briefly, the algorithm obtains tissue-specific gene expression measured in average RPKM (Reads per Kilobase of transcript, per million mapped reads) from 53 tissue from GTEx-V7. Theses gene expression measures are then log2 transformed and winsorized at 50. Finally, gene-property analysis is performed using average expression of genes per tissue type as a gene covariate. Third, we performed cell type enrichment analyses in the brain cortex using FUMA’s web-based CELL TYPE tool. We used the RNA sequencing dataset GSE67835 of human cerebral cortex. This dataset is comprised of 19,749 genes measured in 331 cells from adult human brain samples ^43^. First, a regression analysis was performed in the selected dataset to select significant cell types. Second, a stepwise conditional analysis to distinguish between correlated cell types was performed. These analyses allowed to test the hypothesis that the genes were enriched in a specific cell type.

#### A brain protein quantitative trait loci genetic risk score and eating behaviors and dietary intakes in the Québec Family Study

We computed a brain pQTL genetic risk score in participants of Québec Family Study (NCT03355729) from which we had complete data on macronutrient intake and genetic data (n = 750). Dietary intake was assessed with a 3-day food record on 2 weekdays and 1 weekend day ^44^. A total of 602 participants also completed the three-factor eating questionnaire, as previously described ^18^. Genotyping of participants was performed using the Illumina 610-Quad Chip, as previously described ^45^. A brain pQTL GRS was calculated by including the candidate causal variant prioritized by colocalization analysis for each causal protein directly measured or imputed in QFS [67 independent (LD R2 < 0.01) variants]. Weight for each SNP was calculated using the effect size and standard error of the summary statistics of largest BMI GWAS ^12^. We included as covariates under- and overreporters of energy intake status, as defined using the method described by Huang et al. ^46^. The effect of the brain pQTL GRS on macronutrient intake was assessed with linear regression correcting for age, sex, and overreporter and underreporter status. For comparison, we calculated a second GRS using the 67 BMI lead SNPs from the GWAS by Locke et al., ^6^. Similarly, we used the effect size and standard errors from the Pulit et al. GWAS as weight to construct this top BMI GRS.

#### Shared genetic etiology between brain protein quantitative trait loci and dietary habits in the UK Biobank

We performed multi-trait colocalization analyses including GWAS summary statistics on 53 heritable dietary habits derived from a shortened food frequency questionnaire in participants of the UK Biobank ^19^. Given the correlation between the different items of the questionnaires, we also included the ten first principal components, resulting in 63 dietary habits exposures evaluated ^19^. We first performed colocalization analysis using the same parametrization as described above between all causal proteins and 63 dietary habits exposures. When proteins colocalized with more than one dietary habits, we performed multi-trait colocalization analyses using the same parametrization as described above to evaluate shared genetic etiology across multiple traits.

#### Code Availability

Code was performed in the R V.4.0.0 computing environment using publicly accessible functions from the TwoSampleMR V.0.5.6 https://github.com/MRCIEU/TwoSampleMR, the MendelianRandomization V.0.5.1 https://cran.rproject.org/web/packages/MendelianRandomization/index.html and the data.table V.1.14.0 https://github.com/Rdatatable/data.tablepackages. The coloc package V.5.1.0 https://github.com/chr1swallace/coloc. The hyprcoloc package V.1.0 https://github.com/jrs95/hyprcoloc. The susieR package V.0.11.93 https://github.com/stephenslab/susieR. The tidyverse V.1.3.1 collection of R packages (Wickham et al. 2019), the gwasglue package V.0.0.0.9000 and the gwasvcf V.0.1.0 package was also used for data wrangling.

#### Data Availability

All genome-wide summary statistics used in this study are in the public domain. Data described in the manuscript from the QFS will be made available upon request pending approval from the authors as well as the funding bodies.

## ACKNOWLEDGEMENTS

We would like to thank all study participants as well as all investigators of the studies that were used throughout the course of this investigation. EG holds a master’s research award from the *Fonds de recherche du Québec: Santé* (FRQS). BJA holds a junior scholar award from the FRQS. P.M. is the recipient of the Joseph C. Edwards Foundation granted to Université Laval. CB is partially supported by the John W. Barton Sr. Chair in Genetics and Nutrition.

